# Comparative Effectiveness of BNT162b2 and NVX-CoV2373 Vaccines in Korean Adults

**DOI:** 10.1101/2023.02.18.23286136

**Authors:** Seon Kyeong Park, Young June Choe, Seung Ah Choe, Benjamin John Cowling, Ji Hae Hwang, Ju Hee Lee, Kil Hun Lee, Seonju Yi, Sang Won Lee, Geun-Yong Kwon, Eun Jung Jang, Ryu Kyung Kim, Young-Joon Park

## Abstract

**Background:** Various types of vaccines against SARS-CoV-2 have reduced the burden of coronavirus diseases 2019 (COVID-19) across the world. We conducted an observational study to evaluate the effectiveness of NVX-CoV2373 and BNT162b2 in providing protection in Korean adults.

**Methods:** This study was a retrospective matched cohort study to emulate a target trial of three doses of NVX-CoV2373 (N-N-N) versus three doses of BNT162b2 (B-B-B) vaccines in presumed immune-naive adults. We used data from the Korea COVID-19 Vaccine Effectiveness (K-COVE) cohort, combining all COVID-19 laboratory-confirmed cases and all COVID-19 immunization registry, between February and November 2022. We calculated 40-week risk differences and risk ratios between the two vaccines.

**Results:** A total of 3,019 recipients of NVX-CoV2373 vaccine and 3,027 recipients of BNT162b2 vaccine were eligible for the study. The 40-week risk ratios for recipients of the NVX-CoV2373 vaccine as compared with recipients of the BNT162b2 vaccine were 1.169 (95% CI, 1.015 to 1.347) for laboratory-confirmed SARS-CoV-2 infection, and 0.504 (95% CI, 0.126 to 2.014) for severe SARS-CoV-2 infection. Estimated risk of severe infection was 0.001 events per 1000 persons (95% CI, 0 to 0.003) for the NVX-CoV2373 vaccine and 0.002 events per 1000 persons (95% CI, 0.001 to 0.006) for BNT162b2 vaccine.

**Conclusion:** This study identifies reduced risk of SARS-CoV-2 infection and severe infection after receipt of three doses of either NVX-CoV2373 or BNT162b2 vaccines in Korean adults. Direct, vaccine-conferred protection may be of importance among high risk persons to mitigate from serious clinical outcome from COVID-19.

## Introduction

Various types of vaccines against SARS-CoV-2 have reduced the burden of coronavirus diseases 2019 (COVID-19) across the world [1]. However, global problem with supply and ongoing demand for booster doses of COVID-19 vaccines makes large portion of population under-immunized [2]. Moreover, allergic reaction to certain types of vaccines calls upon the medical needs for other options to prevent from severe COVID-19 [3, 4].

The recombinant protein-based vaccine NVX-CoV2373 has demonstrated high immunogenicity and efficacy against COVID-19 and has been introduced in more than 40 countries [5, 6]. Despite its use in different settings, the comparative effectiveness of NVX-CoV2373 and other COVID-19 vaccines for the prevention of SARS-CoV-2 infection has yet to be assessed.

Since the beginning of COVID-19 pandemic, South Korean government has been operating a centralized public health database that collects all reported COVID-19 cases and their vaccination status [7]. The NVX-CoV2373 vaccine was introduced in South Korea on March 2022, and as of February 2023, more than 300,000 doses have been distributed [8]. With real-time vaccine registry merged with COVID-19 surveillance data, there is an opportunity to assess the comparative effectiveness of NVX-CoV2373 vaccine [9, 10].

Herein, we conducted an observational study in which we evaluated the effectiveness of NVX-CoV2373 and BNT162b2 in providing protection against SARS-CoV-2 infection and severe infection in Korean adults.

## Methods

This study was a retrospective matched cohort study to emulate a target trial of three doses of NVX-CoV2373 (N-N-N) versus three doses of BNT162b2 (B-B-B) vaccines in presumed immune-naive adults. We used data from the Korea COVID-19 Vaccine Effectiveness (K-COVE) cohort, a large-linked database combining all COVID-19 laboratory-confirmed cases and all COVID-19 immunization registry in South Korea. Detailed information about the K-COVE cohort has been published previously [11-13].

In current study, 4,547,432 Korean adults _≥_18 yrs. of age without a previously documented SARS-CoV-2 infection or COVID-19 vaccination, as of February 14, 2022, were initially screened. Eligibility criteria included all Korean nationals aged at least 18 years, no previously documented SARS-CoV-2 infection, and has had three doses of NVX-CoV2373 or BNT162b2 vaccines between February 14 and November 26, 2022 (286 days). Matching criteria included age, sex, geographic region, and health status (immunocompromised vs. not). The detailed case selection flow is shown in Figure 1.

**Figure 1.**
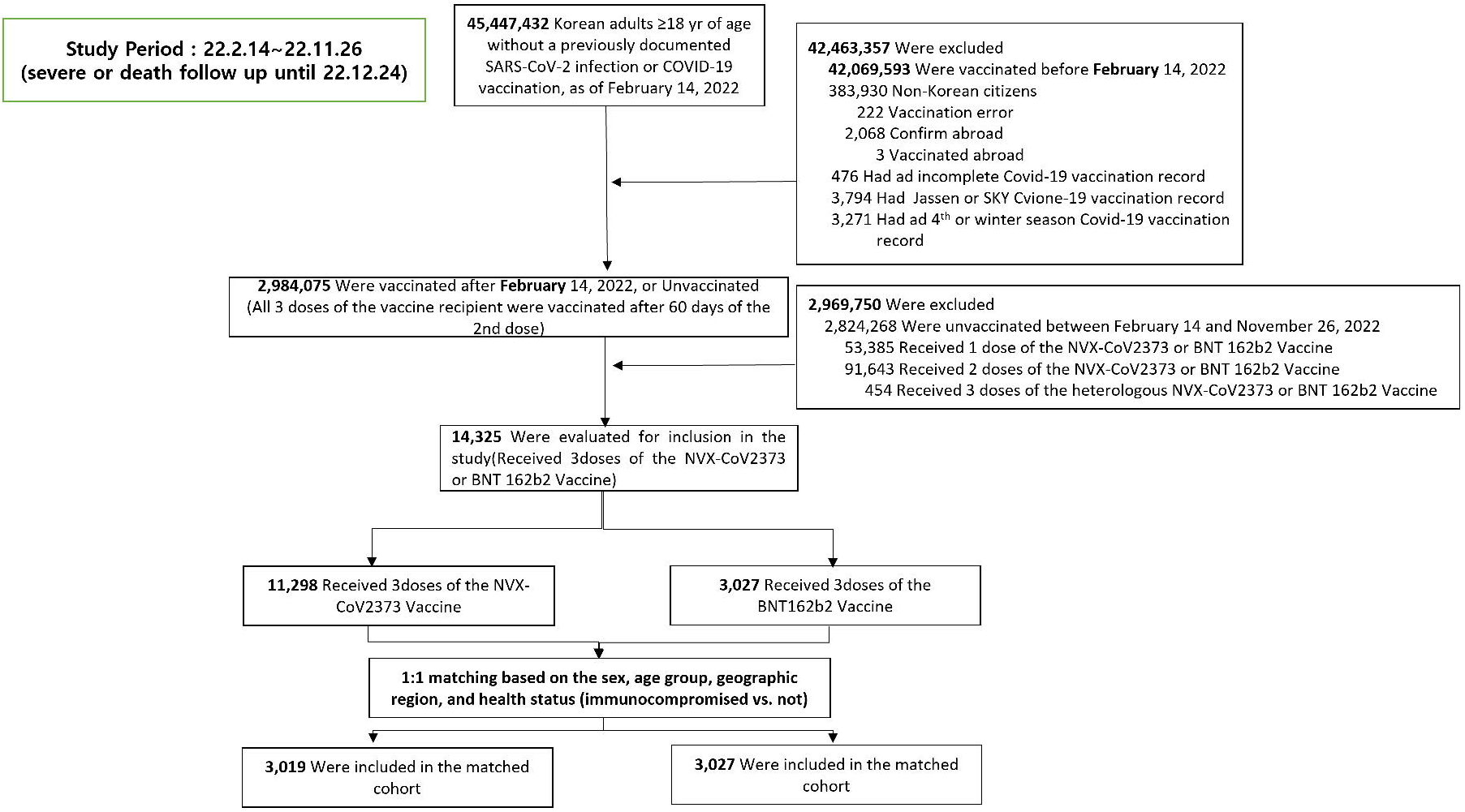
Selection of Persons for the Emulation of a Target Trial Evaluating the Comparative Effectiveness of the NVX-CoV2373 and BNT162b2 Vaccines during a Period Marked by SARS-CoV-2 Omicron-Variant Predominance (February - November, 2022).

**Figure 2.**
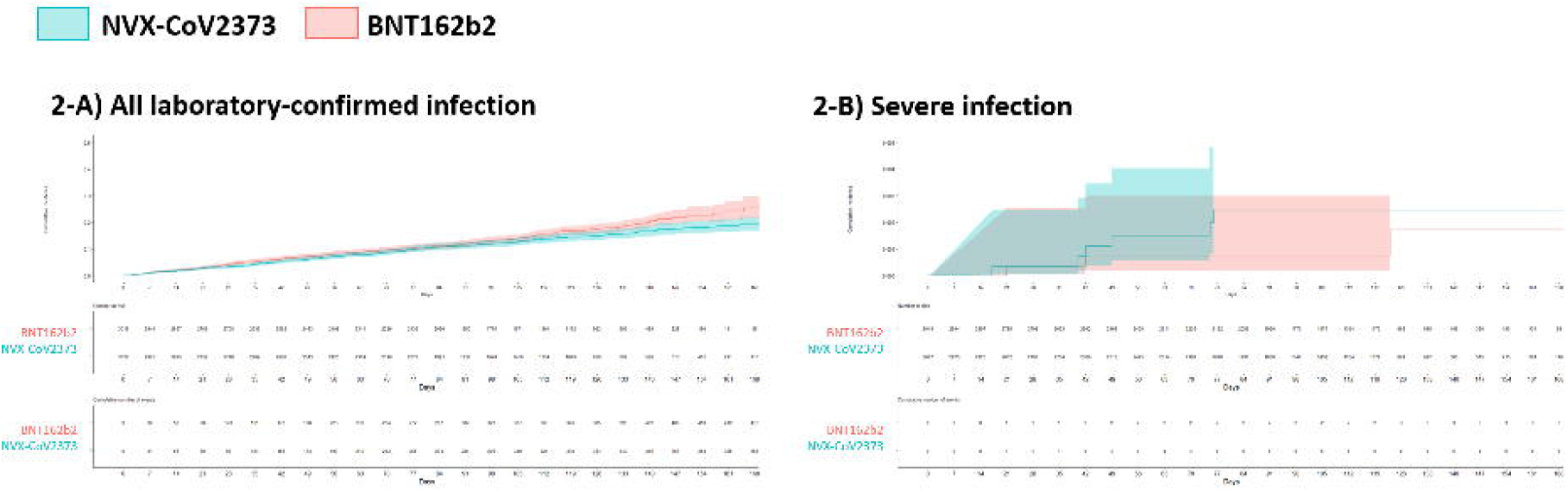
Cumulative Incidence of COVID-19 Outcomes during a Period Marked by SARS-CoV-2 Omicron-Variant Predominance (February-November, 2022).

The interventions of interest were the third-dose vaccination with NVX-CoV2373 vaccine (after receipt of two doses of NVX-CoV2373) or the third-dose of BNT162b2 vaccine (after receipt of two doses of BNT162b2). Between February and November 2022, there was no preferential recommendation issued by the government or academic society between NVX-CoV2373 or BNT162b2 vaccines, and all persons aged 12 years and above were eligible for free vaccination during this period. Per national guideline, both NVX-CoV2373 BNT162b2 vaccine’s schedules were as follows: two doses in interval of 3 weeks; followed by a third dose booster after 3 months, and a fourth dose booster after 4 months. The third dose booster vaccination was recommended for all adults aged 18 years and above; while the forth dose booster vaccination was first recommended to immunocompromised patients (18-49 years) and adults aged 50 years and above, but was opened to all adults who were willing to get vaccinated. Per national guideline, booster series with homologous vaccine type was recommended (i.e., N-N-N or B-B-B).

The two outcomes of interest were (1) laboratory-confirmed SARS-CoV-2 infection, and (2) severe SARS-CoV-2 infection. Laboratory-confirmed SARS-CoV-2 infection include either rapid antigen test (RAT) and polymerase chain reaction (PCR) test, conducted by healthcare professionals. Severe SARS-CoV-2 infection is defined if the patient was treated with high flow oxygen, invasive mechanical ventilation, extracorporeal membrane oxygenation, continuous renal replacement therapy, or death attributable to COVID-19, within 30 days following diagnosis of COVID-19.

For each eligible participant, follow-up started on the day the first dose of vaccine was received and ended on the day of the outcome of interest or the end of the study, whichever occurred first. Observed period was from February 14, 2022, to November 26, 2022 for SARS-CoV-2 infection; and for severe infection, the observed period was extended through December 24, 2022. Supplemental Figure 1 shows the trend of national virus genomic surveillance data suggesting that BA.2, BA.2.3., and BA.5 were the main circulating subvariant during the observed period.

We used the Kaplan–Meier estimator with daily outcome events to compute the risk of the outcome during the period, then calculated 40-week risk differences and risk ratios between the NVX-CoV2373 and BNT162b2 groups. Statistical analyses were conducted using R v.4.02 (R Core Team, Vienna, Austria).

This study was conducted as a legally mandated public health investigation under the authority of the Korean Infectious Diseases Control and Prevention Act. The study was approved by the Korea Disease Control and Prevention Agency Institutional Review Board (IRB No. 2021-12-03-PE-A).

## Results

Among the 14,325 Korean adults who received their third dose of NVX-CoV2373 or BNT162b2 vaccines after February 14, 2022, a total of 3,019 recipients of NVX-CoV2373 vaccine and 3,027 recipients of BNT162b2 vaccine were eligible for the study (Figure 1). The baseline characteristics of matched persons is shown in Table 1. The median age was 54 years in both groups, female-to-male ratio was 55:45, and 0.1%-0.2% were long-term care facility residents, and 7.4%-7.5% were immunocompromised patients. The median follow-up period was 97.19 days for NVX-CoV2373 recipients and 97.61 days for BNT162b2 recipients. Supplementary Table 1 shows the characteristics of unmatched NVX-CoV2373 recipients, and Supplementary Table 2 shows the characteristics of unvaccinated, 1-dose vaccinated, 2-dose vaccinated, and those who received heterologous booster 3-doses.

Over a 40-week follow-up period, there were 769 SARS-CoV-2 laboratory-confirmed infection and of those, nine cases (1.1 %) were classified as severe infection (Table 2). Of the 413 laboratory-confirmed-infection among NVX-CoV2373 recipients, three cases (0.7%) were classified as severe infection; while of the 356 laboratory-confirmed-infection among BNT162b2 recipients, six (1.7%) were classified as severe infection. The 40-week risk ratios for recipients of the NVX-CoV2373 vaccine as compared with recipients of the BNT162b2 vaccine were 1.169 (95% CI, 1.015 to 1.347) for laboratory-confirmed SARS-CoV-2 infection, and 0.504 (95% CI, 0.126 to 2.014) for severe SARS-CoV-2 infection (Table 2). The risk differences (BNT162b2 minus NVX-CoV2373), expressed as events over 40 weeks per 1,000 persons, were 1.214 (95% CI, 1.083 to 1.337) for laboratory-confirmed SARS-CoV-2 infection, and 0.020 (95% CI, 0.007 to 0.044) for severe infection. Supplementary Table 3 shows age-specific risk ratio and risk difference.

Figure 1 shows the cumulative incidence curves for the study outcomes in each vaccine groups. The absolute risks of the outcomes were low in both groups, with estimated risk of all laboratory-confirmed infection was 0.162 events per 1000 persons (95% CI, 0.147 to 0.178) for the NVX-CoV2373 vaccine and 0.140 events per 1000 persons (95% CI, 0.126 to 0.155) for BNT162b2 vaccine. Estimated risk of severe infection was 0.001 events per 1000 persons (95% CI, 0 to 0.003) for the NVX-CoV2373 vaccine and 0.002 events per 1000 persons (95% CI, 0.001 to 0.006) for BNT162b2 vaccine.

## Discussion

The target trial emulation study we performed in the South Korea demonstrated that the three-dose BNT162b2 had lower risk of all laboratory-confirmed infection compared to NVX-CoV2373, however, the risk of severe infection was similarly low in both vaccines. Our finding provides strong support for the current indication and recommendation that NVX-CoV2373 provide robust effectiveness against COVID-19 and its serious outcome. Also, the result is in line with previous meta-analysis that pooled 57 studies that showed vaccine effectiveness of 77.1%-98.0% and 86.3%-93.6% for BNT162b2 and NVX-CoV2373 vaccines against SARS-CoV-2 infection, respectively [14]. An Australian study that compared rates of SARS-CoV-2 Omicron BA.1/2 infection showed an adjusted hazard ratio for SARS-CoV-2 infection of NVX-CoV2373 recipients compared to BNT162b2 vaccine was 1.70 (1.46-1.97) [15]. These findings may be related to NVX-CoV2373 vaccine’s less inducing of spike-specific CD8 T cells compared to mRNA vaccines, as demonstrated in a German immunogenicity study [16].

Despite the difference in protection against all SARS-CoV-2 infection between the two vaccines, our finding suggests a comparable protective effect against severe infection in both NVX-CoV2373 and BNT162b2 vaccines. A recent randomized trial from the U.S. and Mexico showed the vaccine efficacy of NVX-CoV2373 against moderate-to-severe COVID-19 of 100%, which is in line with our finding [17]. Other immunologic data supports the result we present. A comparative immunogenicity study with BNT162b2 or mRNA-1273 showed that the NVX-CoV2373 have strongly induced spike-specific antibodies and the CD4 T-cells equally recognized all tested variants from Alpha to Omicron [16], which is further supported by a data showing NVX-CoV2373 inducing high functional T-cell immunity [18]. Together with the real-world data that we present, these findings suggest a robust evidence that NVX-CoV2373 provides enhanced protection against severe disease caused by SARS-CoV-2.

The observed period was predominated by the Omicron BA.1, BA.2, and BA.4/5 variants, therefore, our finding suggests a net benefit from the predecessor vaccine with original strain in mitigating from serious health outcome from SARS-CoV-2 variant [19, 20]. A study evaluating immunogenicity of fourth homologous dose of NVX-CoV2373 showed an enhanced cross-reactive immunity to SARS-CoV-2 variants, suggesting broader protection against emerging SARS-CoV-2 variants [21]. These findings suggest that under current level of circulation of Omicron BA.5 subvariant and in the context of primary immunization with monovalent COVID-19 vaccines, receipt of booster doses may substantially reduce the disease burden in adult population. The homologous booster dose of NVX-CoV2373 vaccine in a phase 2 trial suggested a robust immunogenicity of IgG geometric mean titers (GMT) increased of more than 4-folds against ancestral SARS-CoV-2 strain [22]. In the current study, we only included those who were vaccinated with all three doses of NVX-CoV2373, however, a further study is needed to assess the effectiveness of heterologous vaccination with other types of vaccines. One study that measured heterologous NVX-CoV2373 booster vaccination showed enhanced cross-reactive immunity against Omicron BA.1/BA.5 subvariants [23].

.This study has limitations. First, our finding does not account for potential changes over time in indirect protection as a result of change in population immunity from natural infection but this should not have a differential effect on recipients of NVX-CoV2373 or BNT162b2. Second, we were unable to examine comorbidities and other unmeasured socioeconomic factors and their impact on choice of vaccination or risk of infection, and our results might therefore suffer from confounding. Lastly, SARS-CoV-2 genotypes in individual breakthrough cases were unknown, which may limit the interpretation of our results. Despite these limitations, a key strength of this study is that it showed NVX-CoV2373 vaccine’s effectiveness in a large scale of population, including population with an increased risk of COVID-19. Long-term follow-up ins planned to compare the results in NVX-CoV2373 recipients with those who received other vaccines.

In conclusion, our study identifies reduced risk of SARS-CoV-2 infection and severe infection after receipt of three doses of either NVX-CoV2373 or BNT162b2 vaccines in Korean adults. Direct, vaccine-conferred protection may be of importance among high risk persons to mitigate from serious clinical outcome from COVID-19. Updated assessments of the vaccine effectiveness of other COVID-19 vaccines are warranted in light of real-world evidence of vaccine direct effects against evolving variants of SARS-CoV-2.

## Data Availability

All data produced in the present study are available upon reasonable request to the authors.

## Acknowledgement

We thank the K-COVE data team for their roles in developing and managing the datasets. We also thank Dr. Mark Thompson and Dr. Muruga Vadivale (both from Novavax Inc.) for advice and feedback in developing this study. Data is not publicly available.

## Author contribution

Conceived and designed the analyses: S.K.P., Y.J.C., Y.J.P., Performed data extraction and analyzed data: S.K.P., H.H., J.H.L., K.H.L., Data curation and project management: S.Y., G.Y.K., E.J.J., R.K.K., Contributed to analysis: Y.J.C., S.A.C., B.J.C., Y.J.P., Wrote the paper: S.K.P., Y.J.C., Y.J.P.

## Funding

None

## Figure legends

Supplementary Figure 1. SARS-CoV-2 subvariants by surveillance week

## Notes

### Competing Interest Statement

The authors have declared no competing interest.

### Funding Statement

This study did not receive any funding.

