## Supplementary material for "Comparative Effectiveness of BNT162b2 and NVX-CoV2373 Vaccines in Korean Adults": Tables

Table 1. Baseline Characteristics of Matched Persons in the Target-Trial Emulation Evaluating the Comparative Effectiveness of the NVX-CoV2373 and BNT162b2 Vaccines during a Period Marked by Omicron-Variant Predominance (February - November 2022)

| Characteristic | NVX-CoV2373 Recipients | | BNT-162b2 Recipients | | Standardized Mean Difference* |
| --- | --- | --- | --- | --- | --- |
|  | (N=3,019) | | (N=3,027) | |  |
| Median age - yr | 54 |  | 54 |  |  |
| Age group - no. (%) |  |  |  |  | 0.004 |
| 0/18-39 yr | 871 | 28.9% | 871 | 28.8% |  |
| 40-59 yr | 876 | 29.0% | 878 | 29.0% |  |
| 60-69 yr | 447 | 14.8% | 452 | 14.9% |  |
| 70-79 yr | 345 | 11.4% | 346 | 11.4% |  |
| ≥80 yr | 480 | 15.9% | 480 | 15.9% |  |
| Sex - no. (%) |  |  |  |  | 0.003 |
| Female | 1,667 | 55.2% | 1,667 | 55.1% |  |
| Male | 1,352 | 44.8% | 1,360 | 44.9% |  |
| Residence |  |  |  |  | 0.002 |
| Metropolitan residence - no. (%) | 1,634 | 54.1% | 1,635 | 54.2% |  |
| Non-metropolitan residence - no. (%) | 1,385 | 45.9% | 1,392 | 45.8% |  |
| Health Status |  |  |  |  | 0.026 |
| LTCF residence - no. (%) | 3 | 0.1% | 6 | 0.2% |  |
| Immunocompromised state - no. (%) | 223 | 7.4% | 228 | 7.5% |  |

*The standardized mean difference is the difference between the number of NVX-Cov2372 recipient and the number of BNT-162b2 Recipients.

**Abbreviations: IQR, interquartile range; LTCF, long-term care facility

Table 2. Estimated Comparative Effectiveness of the NVX-CoV2373 and BNT162b2 Vaccines during a Period Marked by Omicron-Variant Predominance (February- November, 2022).

| Covid-19 Outcome | No. of Events | | 40-Week Risk (95% CI) | | |  | Risk Difference (95% CI) | Risk Ratio (95% CI) |
| --- | --- | --- | --- | --- | --- | --- | --- | --- |
|  | NVX-CoV2373 | BNT162b2 | NVX-CoV2373 | 95% CI | BNT162b2 | 95% CI |  |  |
|  |  |  | events/1,000 persons-day |  | events/1,000 persons-day |  | events/1,000 persons-day | |
| All laboratory-confirmed infection | 413 | 356 | 1.408 | (1.276,1.551) | 1.214 | (1.083,1.337) | 0.204 (0.019,0.388) | 1.169 (1.015,1.347) |
| Severe infection | 3 | 6 | 0.010 | (0.002,0.030) | 0.020 | (0.008,0.044) | -0.010 (-0.030,0.010) | 0.504 (0.126,2.014) |
