## Supplemental Tables for "Comparative Effectiveness of BNT162b2 and NVX-CoV2373 Vaccines in Korean Adults"

Supplementary Table 1. Baseline Characteristics of Unmatched Persons in the Target-Trial Emulation Evaluating the Comparative Effectiveness of the NVX-CoV2373 and BNT162b2 Vaccines during a Period Marked by Omicron-Variant Predominance (February - November 2022)

| Characteristic | NVX-CoV2373 Recipients | | BNT-162b2 Recipients | | Standardized Mean Difference* |
| --- | --- | --- | --- | --- | --- |
|  | (N=11,298) | | (N=3,027) | |  |
| Median age (IQR) - yr | 56 |  | 54 |  | 0.035 |
| Age group - no. (%) |  |  |  |  | 0.225 |
| 18-39 yr | 2,562 | 22.7% | 871 | 28.8% |  |
| 40-59 yr | 3,779 | 33.4% | 878 | 29.0% |  |
| 60-69 yr | 2,299 | 20.3% | 452 | 14.9% |  |
| 70-79 yr | 1,336 | 11.8% | 346 | 11.4% |  |
| ≥80 yr | 1,322 | 11.7% | 480 | 15.9% |  |
| Sex - no. (%) |  |  |  |  | 0.008 |
| Female | 6,177 | 54.7% | 1,667 | 55.1% |  |
| Male | 5,121 | 45.3% | 1,360 | 44.9% |  |
| Residence |  |  |  |  | 0.005 |
| Metropolitan residence - no. (%) | 5,167 | 45.7% | 1,635 | 54.2% |  |
| Non-metropolitan residence - no. (%) | 6,131 | 54.3% | 1,392 |  |  |
| Health Status |  |  |  |  | 0.164 |
| LTCF residence - no. (%) | 11 | 0.1% | 6 | 0.2% |  |
| Immunocompromised state - no. (%) | 430 | 3.8% | 228 | 7.5% |  |

*The standardized mean difference is the difference between the number of NVX-Cov2372 recipient and the number of BNT-162b2 Recipients.

Supplementary Table 2. Baseline Characteristics of the persons unvaccinated, received 1dose or 2 doses of the NVX-CoV2373 or BNT 162b2 Vaccines during a Period Marked by Omicron-Variant Predominance (February - November, 2022)

| Characteristic | Unvaccinated | | 1 Dose vaccination | | 2 Doses vaccination | | 3 Doses vaccination (heterologous vaccination) | |
| --- | --- | --- | --- | --- | --- | --- | --- | --- |
|  | (N=2,824,268) | | (N=53,358) | | (N=91,643) | | (N=454) | |
| Median age (IQR) - yr | 43 | | 38 | | 41 | | 64 | |
| Age group - no. (%) |  |  |  |  |  |  |  |  |
| 0/18-39 yr | 1,154,744 | 40.9% | 28,391 | 53.2% | 42,389 | 46.3% | 85 | 18.7% |
| 40-59 yr | 1,013,261 | 35.9% | 15,254 | 28.6% | 30,887 | 33.7% | 111 | 24.4% |
| 60-69 yr | 306,146 | 10.8% | 3,778 | 7.1% | 8,987 | 9.8% | 75 | 16.5% |
| 70-79 yr | 160,309 | 5.7% | 2,243 | 4.2% | 4,189 | 4.6% | 69 | 15.2% |
| ≥80 yr | 189,808 | 6.7% | 3,719 | 7.0% | 5,191 | 5.7% | 114 | 25.1% |
| Sex - no. (%) |  |  |  |  |  |  |  |  |
| Female | 1,487,666 | 52.7% | 28,798 | 53.9% | 51,073 | 55.7% | 247 | 54.4% |
| Male | 1,336,602 | 47.3% | 24,587 | 46.1% | 40,570 | 44.3% | 207 | 45.6% |
| Residence |  |  |  |  |  |  |  |  |
| Metropolitan residence - no. (%) | 1,486,549 | 52.6% | 25,942 | 48.6% | 44,598 | 48.7% | 202 | 44.5% |
| Non-metropolitan residence - no. (%) | 1,337,719 | 47.4% | 27,443 | 51.4% | 47,045 | 51.3% | 252 | 55.5% |
| Health Status |  |  |  |  |  |  |  |  |
| LTCF residence - no. (%) | 3 | 0.0% | 3 | 0.0% | 1 | 0.0% | 0 | 0.0% |
| Immunocompromised state - no. (%) | 636 | 0.0% | 505 | 0.9% | 1,914 | 2.1% | 78 | 17.2% |

Supplementary Table 3. Estimated Comparative Effectiveness of the NVX-CoV2373 and BNT162b2 Vaccines Based on Age during a Period Marked by Omicron-Variant Predominance (February- November, 2022).

| Covid-19 Outcome | No. of Events/person-days | | | | 40-Week Risk (95% CI) | | | | Risk Difference (95% CI) | Risk Ratio (95% CI) |
| --- | --- | --- | --- | --- | --- | --- | --- | --- | --- | --- |
|  | NVX-CoV2373 | Person-day | BNT162b2 | Person-day | NVX-CoV2373 | 95% CI | BNT162b2 | 95% CI | events/1000 persons-day | |
|  |  |  |  |  | events/1000 persons-day |  | events/1000 persons-day |  |  |  |
| 18-39 yr |  |  |  |  |  |  |  |  |  |  |
| All laboratory-confirmed infection | 150 | 77,316 | 101 | 79,928 | 1.940 | (1.642,2.277) | 1.264 | (1.029,1.535) | 0.676(0.280,1.073) | 1.535(1.193,1.976) |
| Severe infection | 0 | 77,316 | 0 | 79,928 | 0.000 | - | 0.000 | - | 0.000 | - |
| 40-59 yr |  |  |  |  |  |  |  |  |  |  |
| All laboratory-confirmed infection | 88 | 89,531 | 71 | 89,735 | 0.983 | (0.788,1.211) | 0.791 | (0.618,0.998) | 0.192(-0.084,0.467) | 1.225(0.897,1.762) |
| Severe infection | 0 | 89,531 | 2 | 89,735 | 0.000 | - | 0.022 | (0.003,0.081) | -0.022(-0.053,0.009) | - |
| 60-69 yr |  |  |  |  |  |  |  |  |  |  |
| All laboratory-confirmed infection | 53 | 43,868 | 46 | 46,335 | 1.208 | (0.905,1.580) | 0.993 | (0.727,1.324) | 0.215(-0.218,0.649) | 1.217(0.820,1.806) |
| Severe infection | 0 | 43,868 | 1 | 46,335 | 0.000 | - | 0.022 | (0.001,0.120) | -0.022(-0.064,0.021) | - |
| 70-79 yr |  |  |  |  |  |  |  |  |  |  |
| All laboratory-confirmed infection | 49 | 34,328 | 49 | 33,135 | 1.427 | (1.056,1.887) | 1.479 | (1.094,1.955) | -0.051(-0.626,0.524) | 0.965(0.650,1.438) |
| Severe infection | 1 | 34,328 | 0 | 33,135 | 0.029 | (0.001,0.162) | 0.000 | - | 0.027(-0.028,0.086) | - |
| ≥80 yr |  |  |  |  |  |  |  |  |  |  |
| All laboratory-confirmed infection | 73 | 48,194 | 89 | 46,332 | 1.515 | (1.187,1.905) | 1.921 | (1.543,2.364) | -0.406(-0.935,0.122) | 0.789(0.5789,1.074) |
| Severe infection | 2 | 48,194 | 3 | 46,332 | 0.041 | (0.005,0.150) | 0.065 | (0.013,0.189) | -0.023(-0.166,0.070) | 0.641(0.107,3.835) |
