## Supplementary figures and images for "Comparative Effectiveness of BNT162b2 and NVX-CoV2373 Vaccines in Korean Adults"

### Supplemental Figure

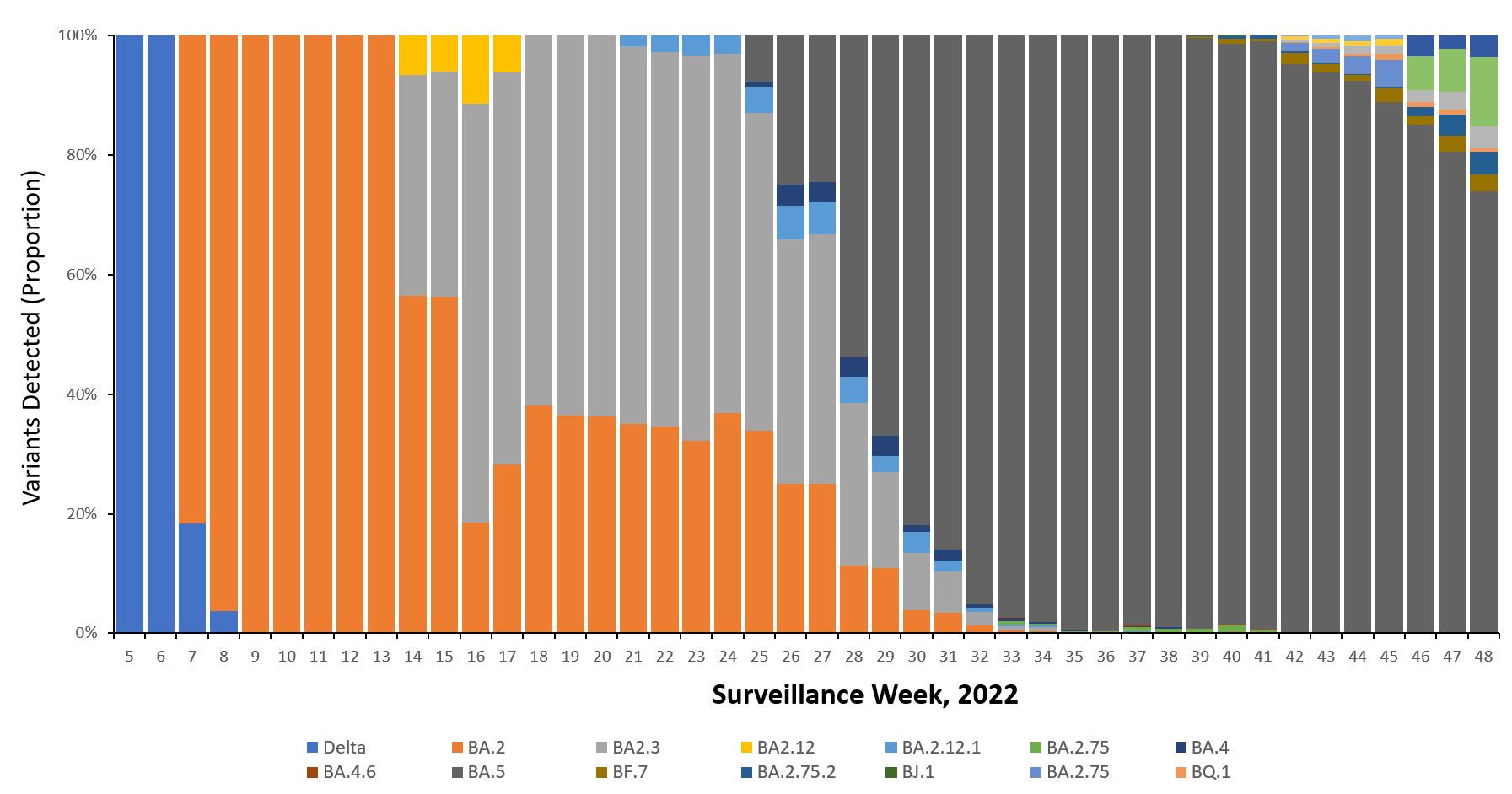
